# Transmission dynamics of COVID-19 in household and community settings in the United Kingdom

**DOI:** 10.1101/2020.08.19.20177188

**Authors:** Jamie Lopez Bernal, Nikolaos Panagiotopoulos, Chloe Byers, Tatiana Garcia Vilaplana, Nicki Boddington, Xu-Sheng Zhang, Andre Charlett, Suzanne Elgohari, Laura Coughlan, Rosie Whillock, Sophie Logan, Hikaru Bolt, Mary Sinnathamby, Louise Letley, Pauline MacDonald, Roberto Vivancos, Oboaghe Edeghere, Charlotte Anderson, Karthik Paranthaman, Simon Cottrell, Jim McMenamin, Maria Zambon, Gavin Dabrera, Mary Ramsay, Vanessa Saliba

## Abstract

**Background:** Households appear to be the highest risk setting for transmission of COVID-19. Large household transmission studies were reported in the early stages of the pandemic in Asia with secondary attack rates ranging from 5–30% but few large scale household transmission studies have been conducted outside of Asia.

**Methods:** A prospective case ascertained study design based on the World Health Organization FFX protocol was undertaken in the UK following the detection of the first case in late January 2020. Household contacts of cases were followed using enhanced surveillance forms to establish whether they developed symptoms of COVID-19, became confirmed cases and their outcomes. Household secondary attack rates and serial intervals were estimated. Individual and household basic reproduction numbers were also estimated. The incubation period was estimated using known point source exposures that resulted in secondary cases.

**Results:** A total of 233 households with two or more people were included with a total of 472 contacts. The overall household SAR was 37% (95% CI 31–43%) with a mean serial interval of 4.67 days, an R_0_ of 1.85 and a household reproduction number of 2.33. We find lower secondary attack rates in larger households. SARs were highest when the primary case was a child. We estimate a mean incubation period of around 4.5 days.

**Conclusions:** High rates of household transmission of COVID-19 were found in the UK emphasising the need for preventative measures in this setting. Careful monitoring of schools reopening is needed to monitor transmission from children.

## Introduction

As of the end of July 2020, over 17 million cases of COVID-19 have been reported globally with over 660,000 deaths (1). The causative agent, SARS-CoV-2, is primarily transmitted through the droplet and contact routes though aerosol and faecal transmission may also contribute (2, 3).

Investigations of household transmission dynamics have been reported in China and other countries in Asia that experienced early cases (4–12). Households appear to be the highest risk setting for transmission with reported secondary attack rates (SARs) in household contacts ranging from 5% to 30% (4–10). Rates of symptomatic infection increase with age and risk factors for more severe disease include age, male sex and a range of comorbidities (13, 14). Other than setting, risk factors for onwards transmission have not been well described. As countries move from broad social distancing measures to more targeted approaches, a detailed understanding of risk factors for transmission is increasingly important.

In the UK the first cases of COVID-19 were reported in late January, the number of cases rapidly increased from March before plateauing, then declining after social distancing measures were introduced (13). We followed up the first few hundred (FF100) cases of COVID-19 in the UK and their household contacts. We have previously reported on the characteristics and outcomes of the cases(13). Here we describe the transmission dynamics and risk factors for transmission and acquisition of symptomatic infection.

## Methods

### Study design

We used a prospective case ascertained study design based on the World Health Organization FFX protocol (15).

### Ascertainment of cases and contacts

The case ascertainment has been described in detail elsewhere (13). Briefly, in the early stages of the pandemic all PCR positive cases who met the case definition were followed up using enhanced surveillance forms on identification and after 14 days. This was later restricted to indigenous cases only. Cases were recruited from February to March 2020.

Close contacts of confirmed cases were identified by the local Health Protection Team (HPT). Those considered at greatest risk, including household contacts, others with direct face to face contact and healthcare workers who had not worn recommended PPE were actively followed up on a daily basis for 14 days and asked about relevant symptoms. Household contacts were defined as those living or spending significant time in the same household. Other contacts not classified as close contacts were provided with health advice and advised to contact the HPT if they developed relevant symptoms. HPTs completed enhanced surveillance questionnaires to collect details from cases on symptoms, medical history, details of the exposure, outcome and any virological tests (supplementary appendix 1). A team of trained staff (health protection practitioners, nurses, doctors and field epidemiologists) proactively followed up all household contacts of confirmed cases 14 days or more after symptom onset in the index case using telephone interviews to assess subsequent development of any symptoms and final outcomes (supplementary appendix 2). Contacts who developed symptoms compatible with COVID-19 were offered PCR testing as per national guidance (13).

If cases or contacts were unable to be contacted by phone after at least two attempts, or if health protection teams had recorded a request for no further contact, they were classified as lost to follow-up.

Details on other non-household community contacts were obtained through the HPZone public health management system. Community contacts with any point source exposures were included where there were no other suspected exposures and complete information was available on the timing of the exposure and symptom onset in the contact. Healthcare workers, returning travellers and airplane exposures were excluded. A detailed dataset was also maintained with information on community exposures and outcomes among all possible contacts of the first 6 cases.

## Analysis

### Household analysis

Confirmed cases were those that tested positive for SARS-CoV-2 on PCR. Probable cases were those with fever, anosmia or respiratory symptoms. Those who had other unrelated or pre-existing illnesses were excluded. FF100 cases and household contacts were reclassified using date of symptom onset, to identify any primary cases that were initially recruited as contacts, and when secondary cases were due to household transmission. Households with two or more household members were included. The probable or confirmed case within the household with the earliest onset date was defined to be the primary household case. When two or more household members had the same earliest symptom onset dates these were defined as co-primary cases, as was any case with symptom onset the day after a primary case.

All other subjects with later symptom onset dates were defined as a secondary case, apart from those that had symptom onset dates greater that 14 days after the primary case.

Initial descriptive analyses were performed to explore the characteristics of the contacts. SARs and odds ratios for secondary transmission were estimated for a range of factors using univariate analyses and multivariate mixed effects logistic regression models with a random intercept for households. The following potential explanatory variables were examined: household size; characteristics of the contact, including: gender and age group; characteristics of the index case, including: gender, age, whether the case was admitted to hospital, and whether the symptoms included coughing or sneezing. Adjusted marginal SARs were estimated for each explanatory variable. Presence or absence of comorbidities among the primary case and contacts were explored as interaction terms.

For the primary analyses co-primaries were excluded and SARs were based on confirmed and probable secondary cases. Three sensitivity analyses were undertaken: 1) with co-primaries included; 1) restricted to laboratory confirmed secondary cases only; 2) with probable confirmed and possible secondary cases, the latter included those who developed any non-respiratory symptoms (e.g. nausea, fatigue, joint aches) within 14 days of exposure.

Serial interval was defined as time from onset of first symptom in the primary cases to time of onset of first symptom in the secondary case with a cut off of 14 days. The same explanatory variables as in the SAR analysis were considered. A lag factor was added to account for cases who were not present the household at the time the symptoms of the corresponding index cases started and adjustment was also made for the number of cases in the household at the time of first exposure. Individual variables were initially explored using Kaplan-Meier estimates of the survival function. Survival regression was then undertaken using the best fitting of the Log-normal, Gamma or Weibull distributions.

Individual basic reproduction number(R_0_) is estimated using the exponential growth model described by Wallinga and Lipsitch (2007) and the renewal equation model described by Fraser (2007) with adjustment for the contribution of imported cases (16, 17). We used the approach described by Fraser (2007) to estimate the household reproduction number (defined as the number of households infected by each infected household) (17). For estimates of reproduction number analyses were restricted to cases from the very early stages of the pandemic when all identified cases were included in the FF100.

### Community contacts

The median incubation period was estimated for probable secondary cases and confirmed secondary cases who had a point source exposure. Exposures before the onset date in the index case were excluded. The timing of exposure among these cases was compared to timing of exposure among contacts that did not develop symptoms. Healthcare workers, returning travellers, airplane exposures and those who had contact with multiple cases were excluded from this analysis.

### Ethics

This was an observational surveillance system carried out under the permissions granted under Regulation 3 of The Health Service (Control of Patient Information) Regulations 2020 and under Section 251 of the NHS Act 2006.

## Results

### Characteristics of cases

The initial FF100 dataset consisted of 379 confirmed COVID-19 cases, 357 from England, 19 from Scotland and three from Wales, who developed symptoms between 24^th^ January 2020 and 13^th^ March 2020. Of the cases 199 were imported, 92 were secondary and 88 indigenous. There were slightly more males (56.7%) than females among the UK FF100 cases. Cases had a mean age of 47.7 years (standard deviation (SD) 17.4) and ranged between 11 months and 94 years. We have previously reported details of these cases and their outcomes (13).

### Recruitment and follow-up of households

After reclassification, there were 365 primary/co-primary cases residing in 329 homes. In 96 households, the case was the only recorded resident. The remaining 269 primary/co-primary cases, resided in 233 homes. 32 households had two co-primary cases and two households had three co-primary cases. In 10 households the primary/co-primary case was under 19 years old. 472 household contacts were identified, of these 32 (6.8%) were lost to follow-up, however 11 were linked to testing data (Figure 1). 135 household contacts developed either cough, fever or anosmia. Among those with tested after symptom onset with complete information on onset and test date the mean time from onset of symptoms to testing of these contacts was 2.9 days.

**Figure 1:**
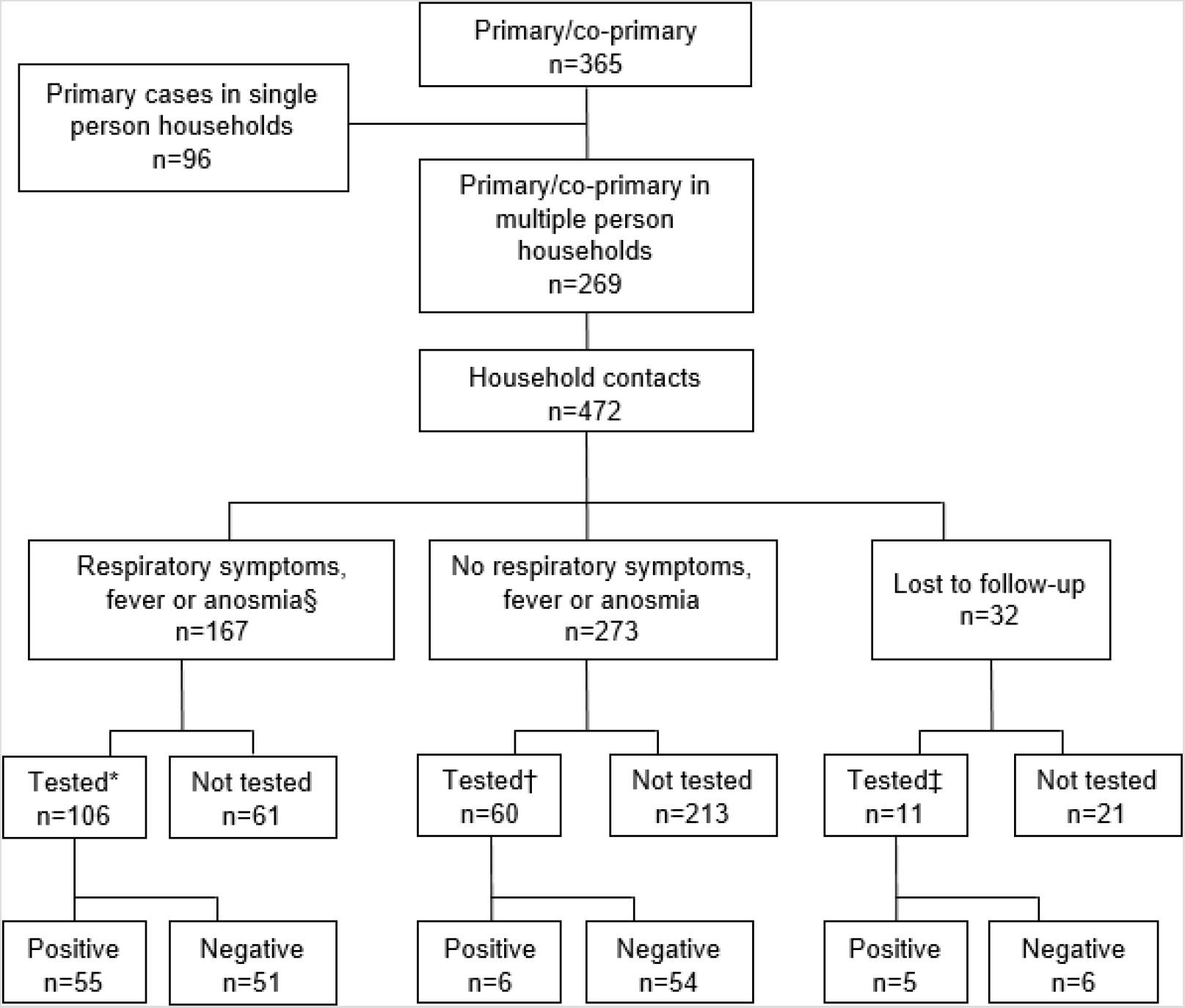
Flowchart of COVID-19 case-patients and household contacts, including contacts with any respiratory symptoms, and contacts with at least one of cough, fever or anosmia, contacts from whom specimens were collected and RT-PCR result, United Kingdom, 2020. §16 persons had onset of symptoms > 2 weeks after onset date in the primary case. *9 persons had specimen date (or laboratory result date if specimen date not known) > 2 weeks after primary case-patient symptoms onset and 4 had a positive test result. †2 persons (neither positive) had specimen date (or laboratory result date if specimen date not known) > 2 weeks after primary case-patient symptoms onset. ‡1 person (not positive) had laboratory result date > 2 weeks after primary case-patient symptoms onset.

### Household contact characteristics

Characteristics of the household contacts are shown in Table 1. Household size ranged from 2 to 7 people. The age of household contacts ranged from 3 months to 84 years, with a mean age of 29.7 years (SD 19.9 years) and 241 (51.1%) were female. Comorbidity data was wholly or partially present for 437 household contacts, 60 (13.7%) of whom had an underlying health condition and 7 (1.6%) of whom had multimorbidity. The most frequent conditions were asthma and other respiratory disease.

**Table 1:** Characteristics of households and household contacts

|  | Non-cases* | Probable<br>secondary cases | Confirmed SARS-CoV-2 | All |
| --- | --- | --- | --- | --- |
| N | 311<br>n(%) | 96<br>n(%) | 65<br>n(%) | 472<br>n(%) |
| Lost to follow-up |  |  |  |  |
| Yes | 24 (7.7) | 3 (3.1) | 5 (7.7) | 32 (6.8) |
| No | 287 (92.3) | 93 (96.9) | 60 (92.3) | 440 (93.2) |
| Household size |  |  |  |  |
| 2 | 39 (12.5) | 17 (17.7) | 21 (32.3) | 77 (16.3) |
| 3 | 63 (20.3) | 25 (26.0) | 18 (27.7) | 106 (22.5) |
| 4 | 104 (33.4) | 30 (31.2) | 18 (27.7) | 152 (32.2) |
| 5 | 61 (19.6) | 18 (18.8) | 7 (10.8) | 86 (18.2) |
| 6 | 22 (7.1) | 4 (4.2) | 1 (1.5) | 27 (5.7) |
| 7 | 22 (7.1) | 2 (2.1) | 0 (0.0) | 24 (5.1) |
| Age |  |  |  |  |
| <19 | 133 (42.8) | 33 (34.4) | 9 (13.8) | 175 (37.1) |
| 19-64 | 166 (53.4) | 63 (65.6) | 50 (76.9) | 279 (59.1) |
| ≥65 | 12 (3.9) | 0 (0.0) | 6 (9.2) | 18 (3.8) |
| Sex |  |  |  |  |
| Female | 161 (51.8) | 46 (47.9) | 34 (52.3) | 241 (51.1) |
| Male | 150 (48.2) | 50 (52.1) | 31 (47.7) | 231 (48.9) |
| Comorbidities |  |  |  |  |
| Any comorbidity | 30 (9.6) | 13 (13.5) | 17 (26.2) | 60 (12.7) |
| Asthma requiring<br>medication | 13 (4.2) | 2 (2.1) | 10 (15.4) | 25 (5.3) |
| Respiratory disease<br>excluding asthma | 5 (1.6) | 3 (3.1) | 2 (3.1) | 10 (2.1) |
| Diabetes (%) | 4 (1.3) | 2 (2.1) | 1 (1.5) | 7 (1.5) |
| Heart disease | 5 (1.6) | 1 (1.0) | 1 (1.5) | 7 (1.5) |
| Immunodeficiency<br>(%) | 4 (1.3) | 1 (1.0) | 1 (1.5) | 6 (1.3) |
| Malignancy | 0 (0.0) | 3 (3.1) | 2 (3.1) | 5 (1.1) |
| Neurological<br>disease (%) | 2 (0.6) | 1 (1.0) | 2 (3.1) | 5 (1.1) |
| Kidney disease | 2 (0.6) | 1 (1.0) | 1 (1.5) | 4 (0.8) |
| Unknown | 29 (9.3) | 3 (3.1) | 3 (4.6) | 35 (7.4) |

\*Non-cases: includes household contacts with no respiratory symptoms, contacts who developed respiratory symptoms >14 days after symptom onset in the primary case, and contacts with unrelated prior illnesses (identified through review of household symptomology dates and HPZone case notes)
\*\*Outcome provided as a percentage of contacts with completed follow-up

Among contacts with complete follow-up the most common symptom was cough (26.1%), followed by fatigue (20.2%) and headache (19.5%) (supplementary figure 1). A response for anosmia was present for 287 contacts, of these 30 (10.5%) experienced anosmia. Of the contacts who developed fever, cough or anosmia 68.1% (92/135) were tested, with 54.3% (50/92) testing positive. Within the follow-up period 3.6% (16/440) of contacts were hospitalised, with a median duration of stay of 3.5 days (IQR 2–9.5 days), all hospitalised contacts tested positive for SARS-CoV-2. None of the contacts with complete follow-up died during the study period.

### Household transmission dynamics

#### SARs

The household secondary attack rate (SAR) was 37% (95% CI 31–43%) including both confirmed or probable secondary cases. If restricted to confirmed secondary cases only the SAR was 16% (95% CI 11–20%) and when possible, probable and confirmed secondary cases were included the SAR was 43% (95% CI 0.37–0.49).

Unadjusted SARs odds ratios of probable and confirmed secondary cases by a range of explanatory variables are shown in Table 2 and the multivariate analysis is shown in Table 3. In both the univariate and multivariate analyses, there was an inverse relationship between household size and SAR, with the highest SAR in households with 2 people (adjusted: 0.48, 95%CI 0.35–0.60) and the lowest in households of 5 or more (0.22, 95%CI 0.12–0.32). There were no significant effects of gender or presence of comorbidities in either primary case or contacts nor of presence of cough or sneezing as a symptom in the primary case. SARs were lowest in contacts aged under 18 years or 65 years and over, however these effects were not significant. SARs were highest where the primary case was aged < 18 years with a significantly higher odds of secondary infection (OR 61, 95% CI 3.3–1133) however there were only 3 households with no coprimaries and a primary case aged under 18 years and there is a lot of uncertainty in this finding. Where the primary case was admitted to hospital there was a significantly lower odds of secondary infection in the household (OR 0.5, 95% CI 0.2–0.8).

**Table 2:**
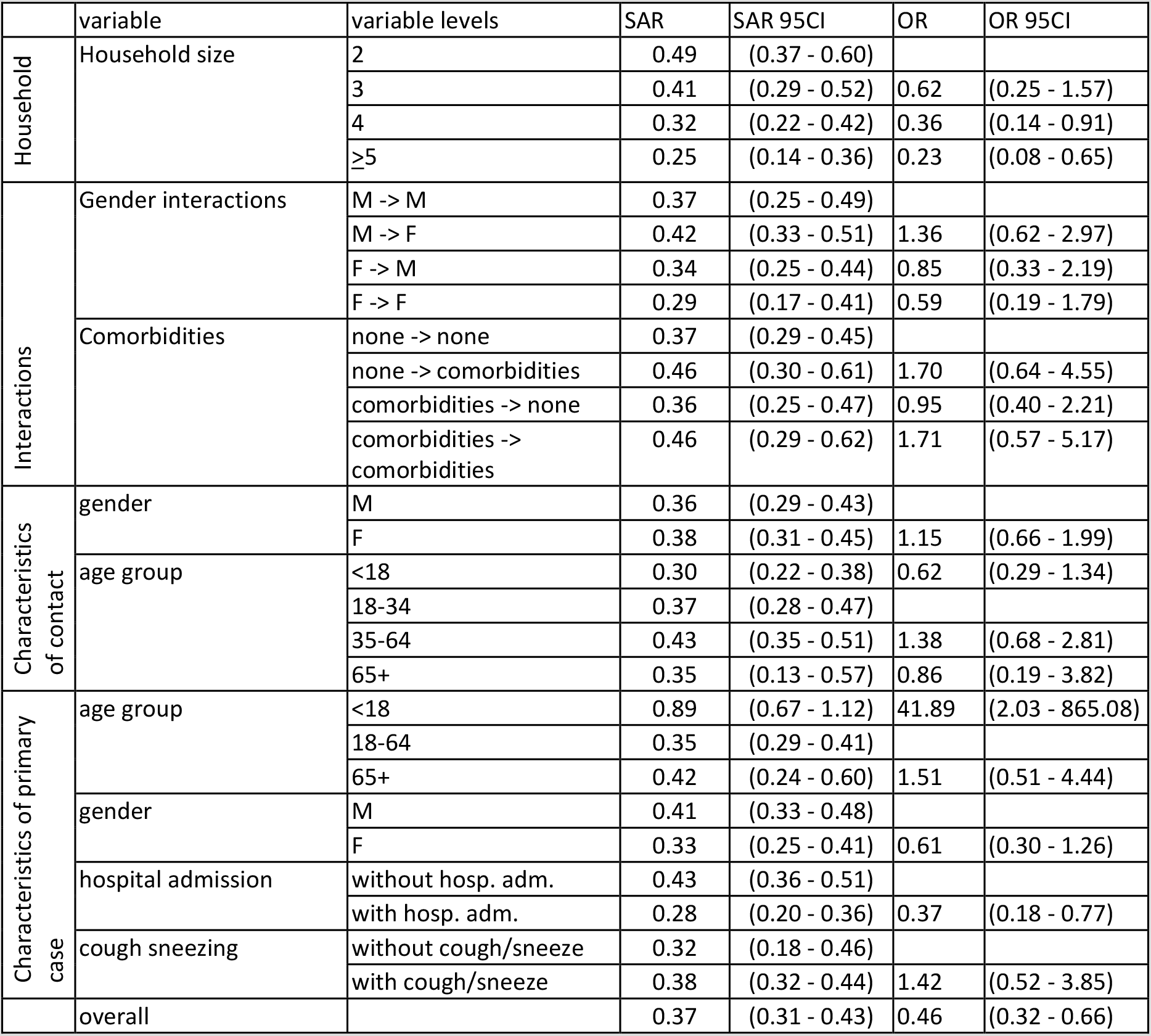
Unadjusted secondary attack rates and odds ratios for secondary infection (probable and confirmed secondary cases)

|  | variable | variable levels | SAR | SAR 95CI | OR | OR 95CI |
| --- | --- | --- | --- | --- | --- | --- |
| Household | Household size | 2 | 0.49 | (0.37 - 0.60) |  |  |
|  |  | 3 | 0.41 | (0.29 - 0.52) | 0.62 | (0.25 - 1.57) |
|  |  | 4 | 0.32 | (0.22 - 0.42) | 0.36 | (0.14 - 0.91) |
|  |  | ≥5 | 0.25 | (0.14 - 0.36) | 0.23 | (0.08 - 0.65) |
| Interactions | Gender interactions | M -> M | 0.37 | (0.25 - 0.49) |  |  |
|  |  | M -> F | 0.42 | (0.33 - 0.51) | 1.36 | (0.62 - 2.97) |
|  |  | F -> M | 0.34 | (0.25 - 0.44) | 0.85 | (0.33 - 2.19) |
|  |  | F -> F | 0.29 | (0.17 - 0.41) | 0.59 | (0.19 - 1.79) |
|  | Comorbidities | none -> none | 0.37 | (0.29 - 0.45) |  |  |
|  |  | none -> comorbidities | 0.46 | (0.30 - 0.61) | 1.70 | (0.64 - 4.55) |
|  |  | comorbidities -> none | 0.36 | (0.25 - 0.47) | 0.95 | (0.40 - 2.21) |
|  |  | comorbidities -> comorbidities | 0.46 | (0.29 - 0.62) | 1.71 | (0.57 - 5.17) |
| Characteristics of contact | gender | M | 0.36 | (0.29 - 0.43) |  |  |
|  |  | F | 0.38 | (0.31 - 0.45) | 1.15 | (0.66 - 1.99) |
|  | age group | <18 | 0.30 | (0.22 - 0.38) | 0.62 | (0.29 - 1.34) |
|  |  | 18-34 | 0.37 | (0.28 - 0.47) |  |  |
|  |  | 35-64 | 0.43 | (0.35 - 0.51) | 1.38 | (0.68 - 2.81) |
|  |  | 65+ | 0.35 | (0.13 - 0.57) | 0.86 | (0.19 - 3.82) |
| Characteristics of primary case | age group | <18 | 0.89 | (0.67 - 1.12) | 41.89 | (2.03 - 865.08) |
|  |  | 18-64 | 0.35 | (0.29 - 0.41) |  |  |
|  |  | 65+ | 0.42 | (0.24 - 0.60) | 1.51 | (0.51 - 4.44) |
|  | gender | M | 0.41 | (0.33 - 0.48) |  |  |
|  |  | F | 0.33 | (0.25 - 0.41) | 0.61 | (0.30 - 1.26) |
|  | hospital admission | without hosp. adm. | 0.43 | (0.36 - 0.51) |  |  |
|  |  | with hosp. adm. | 0.28 | (0.20 - 0.36) | 0.37 | (0.18 - 0.77) |
|  | cough sneezing | without cough/sneeze | 0.32 | (0.18 - 0.46) |  |  |
|  |  | with cough/sneeze | 0.38 | (0.32 - 0.44) | 1.42 | (0.52 - 3.85) |
|  | overall |  | 0.37 | (0.31 - 0.43) | 0.46 | (0.32 - 0.66) |

**Table 3:** Adjusted secondary attack rates and odds ratios for secondary infection (probable and confirmed secondary cases)

|  | Variable | levels | SAR | 95% CI |  | OR | 95% CI |  |
| --- | --- | --- | --- | --- | --- | --- | --- | --- |
| Household | Household size | 2 | 0.48 | 0.35 | 0.6 | 1 |  |  |
|  |  | 3 | 0.4 | 0.29 | 0.52 | 0.67 | 0.27 | 1.6 |
|  |  | 4 | 0.33 | 0.23 | 0.44 | 0.46 | 0.18 | 1.1 |
|  |  | ≥5 | 0.22 | 0.12 | 0.32 | 0.22 | 0.078 | 0.64 |
| Characteristics of contact | Gender | Male | 0.36 | 0.28 | 0.43 | 1 |  |  |
|  |  | Female | 0.32 | 0.24 | 0.39 | 0.8 | 0.44 | 1.5 |
|  | Age group | <18 | 0.29 | 0.2 | 0.38 | 0.73 | 0.34 | 1.6 |
|  |  | 18-34 | 0.34 | 0.24 | 0.44 | 1 |  |  |
|  |  | 35-64 | 0.39 | 0.3 | 0.48 | 1.3 | 0.66 | 2.6 |
|  |  | 65+ | 0.26 | 0.021 | 0.51 | 0.62 | 0.12 | 3.3 |
| Characteristics of primary case | Gender | Male | 0.38 | 0.3 | 0.46 | 1 |  |  |
|  |  | Female | 0.29 | 0.21 | 0.37 | 0.6 | 0.3 | 1.2 |
|  | Age group | <18 | 0.92 | 0.75 | 1.1 | 61 | 3.3 | 1133 |
|  |  | 18-64 | 0.31 | 0.25 | 0.37 | 1 |  |  |
|  |  | 65+ | 0.38 | 0.16 | 0.59 | 1.4 | 0.41 | 5.1 |
|  | Hospital admission | without hospital adm. | 0.4 | 0.33 | 0.48 | 1 |  |  |
|  |  | with hospital adm. | 0.25 | 0.17 | 0.33 | 0.4 | 0.2 | 0.8 |
|  | Cough or sneezing | no cough/sneeze | 0.29 | 0.15 | 0.43 | 1 |  |  |
|  |  | cough/sneeze | 0.34 | 0.28 | 0.41 | 1.4 | 0.54 | 3.4 |

When co-primaries were included in the analysis, results were broadly similar, this increases the number of households with children as a primary and the odds of secondary infection remains significant (OR 8, 95% CI 1.3–49) (supplementary table 1). In the analysis restricted to laboratory confirmed secondary cases, there is a significantly lower odds of secondary infection in contacts aged < 18 years (OR 0.22; 95% CI 0.01–0.87) and the higher odds of secondary infection where the primary case is aged < 18 years remains (OR 22, 95% CI 1–464) (supplementary table 2).

#### Serial interval in households

The Weibull distribution provided the best fit for the univariate survival analysis and gave a mean serial interval of 4.67 days (further details in supplementary figure 2 and supplementary table 3). In the multivariate analysis, explanatory variables that were associated with a shorter serial interval included the primary case experiencing cough as a symptom and the primary case being an imported case (Table 4). There were non-linear relationships between both age of index case and age of household contact and serial interval with shorter serial intervals if the index case was a child or an older adult compared to working age adults and a longer serial interval among household contacts who were children or older adults compared to working age adults (supplementary figure 3). Crude serial intervals and modelled effect of age as a continuous variable are provided in supplementary table 4 and supplementary figure 2 respectively.

**Table 4:**
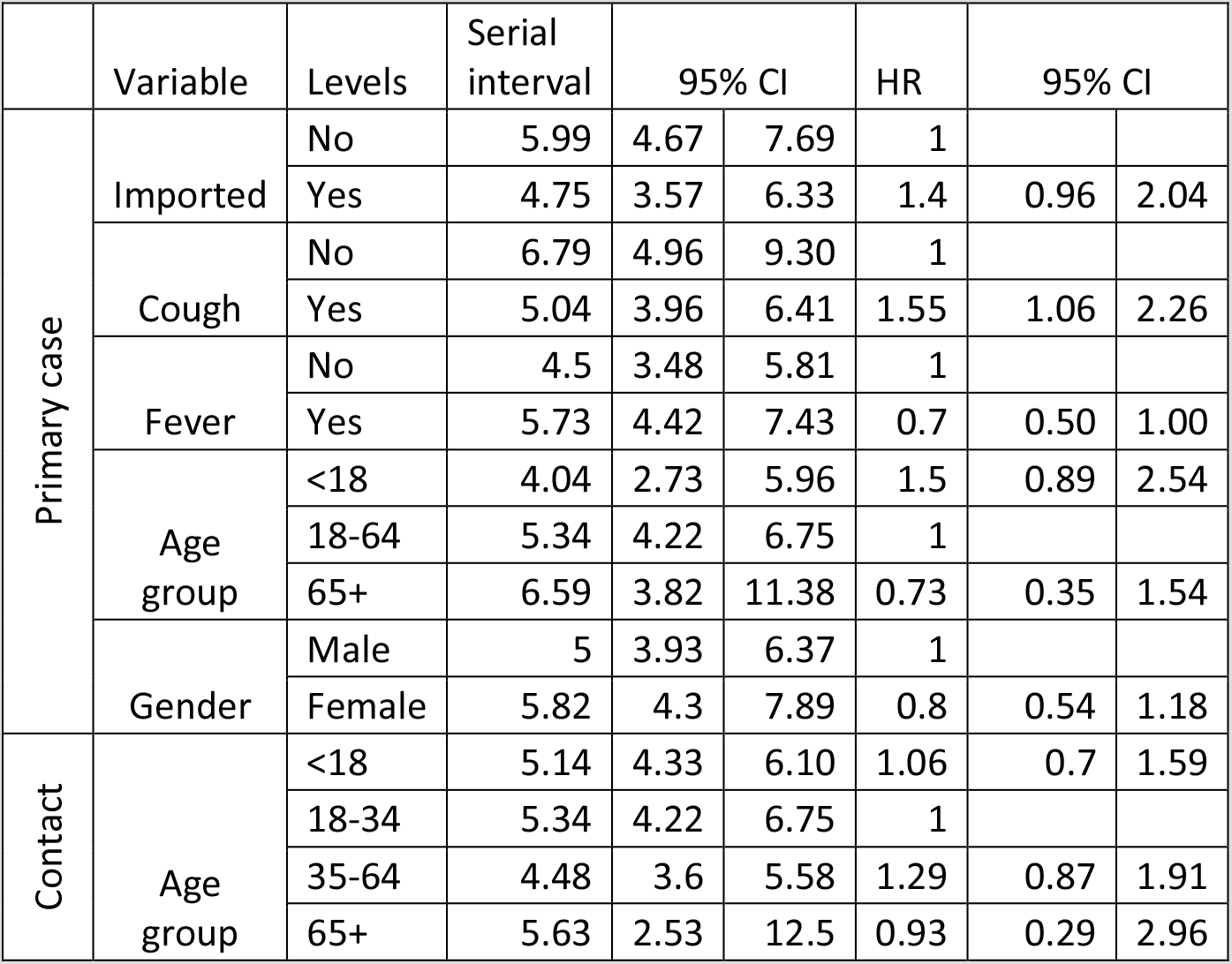
Adjusted serial intervals using marginal means and hazard ratios

#### Basic and household reproduction number

Using the approach by Wallinga and Lipsitch (2007), and based on the serial interval above, we obtained an estimate of R_0_: 3.67 (95%CI: 3.22–3.98). Using the renewal equation to take into account the contribution of imported cases, individual R_0_ in the early stages of the pandemic in the UK is estimated at 1.85 (95% CI: 1.20–3.42). Applying the household transmission model to the household data, we found that the average total number of cases in an infected household is 1.67. The household reproduction number from the same models is estimated at 2.33 (95%CI 1.30–4.89).

### Community contacts

45 confirmed or probable secondary cases were identified that had a point source exposure (exposure window of maximum one day) to a primary case in the FF100 dataset, of these 12 were laboratory confirmed secondary cases. The median incubation period for confirmed and probable cases with a point source exposure was 4.51 days (SD 2.66), for confirmed secondary cases alone it was 4.77 days (SD 2.34) (Table 5). Probable and confirmed secondary cases were exposed a mean of 2.37 days (SD 3.36) after symptom onset in the index case, ranging from 0–14 days. Restricting to confirmed secondary cases alone, exposure was mean 1.33 days (SD 1.61) after symptom onset in the primary case, ranging from 0–5 days). This compares to 2.71 days among contacts who didn’t go on to become a case. Further details of the timing of onset and exposure for the confirmed secondary cases are shown in supplementary figure 4.

**Table 5:** Summary of incubation period and timing of exposure in relation to primary case symptom onset for contacts with a point source exposure.

| Status of contact | Number of contacts included | Incubation period (days) |  |  |  |  | Timing of exposure after symptom onset of primary case (days) |  |  |  |  |
| --- | --- | --- | --- | --- | --- | --- | --- | --- | --- | --- | --- |
|  |  | Mean | SD | Median | IQR | Range | Mean | SD | Median | IQR | Range |
| Probable and confirmed secondary cases | 45* | 4.51 | 2.66 | 4 | 2 - 7 | 0 - 11 | 2.37 | 3.36 | 1 | 0 - 4 | 0 - 14 |
| Confirmed secondary cases | 12 | 4.75 | 2.34 | 4 | 3.75 - 5 | 2 - 11 | 1.33 | 1.61 | 1 | 0 - 1.25 | 0 - 5 |
| Did not develop symptoms | 241 | - | - | - | - | - | 2.71 | 2.74 | 2 | 0 - 5 | 0 - 9 |

\*4 excluded from timing of exposure analysis due to no onset date available for primary case

## Discussion

In the UK, prior to the implementation of social and physical distancing measures, we estimate an overall household SAR of 37%, a serial interval of 4.67 days, an R_0_ of 1.85 and a household reproduction number of 2.33. We find lower secondary attack rates in larger households. There is some suggestion that where the primary case is a child, household SARs are higher and the serial interval is shorter. Conversely serial intervals were longer if the household contact was a child or an older adult. Using point source exposures we estimate a mean incubation period of around 4.5 days.

Our estimated household SAR in the UK is greater than that reported in China, Taiwan and South Korea estimated household SARs ranging from 5% to 30% (4–10). Making comparisons across studies is challenging due to differences in follow-up, symptom ascertainment or testing of contacts, however, the higher household SAR in the UK could reflect differences in isolation and infection control measures taken to reduce spread. In the UK, individuals meeting the case definition were advised to minimise contact with others in the household, wash hands regularly and cover coughs and sneezes. This is broadly similar to advice issued elsewhere, though more stringent advice on quarantine within the household and wearing masks was in place in some areas, and cases were taken out of the household and placed in isolation facilities (4, 8). It is also possible that timing of the diagnosis of secondary cases was more delayed in the very earliest stages of the pandemic in China and other countries that experienced early cases, when less was understood about the disease. Our serial interval estimate is broadly similar to previous estimates that range from 4.0 to 6.3 days.(4, 18–20).

This high estimated R_0_ using the Wallinga and Libsitch (2007) approach is because the method has neglected the contribution of cases that continuously imported from abroad to transmission dynamics in UK. After adjusting for the contribution of imported cases, the R_0_ is lower than existing estimates obtained for the early stage in China, though confidence intervals overlap (21–24).

We estimated a mean incubation period of 4.51–4.75 days, slightly lower than previous estimates which range from 5.5 to 6.4 days (25–27). Previous estimates have been based on estimated distributions using earliest and latest exposure period. In our analysis we restricted to those with unique point source exposures to allow us to precisely estimate exposure date. The incubation period ranged from 2–11 days for confirmed secondary cases and 0–11 days for probable and confirmed combined, suggesting that current advice around isolation of contacts for 14 days after exposure is appropriate. The mean time from onset in the primary case to exposure among confirmed secondary cases was 1.33 days, suggesting that cases are most infectious soon after symptom onset. Though it should be noted that, at the time, contact tracing from the time of symptom onset in the index case, not before symptom onset.

A systematic review and meta-analysis by Viner et al (2020) examined susceptibility of children to SARS-CoV-2 and their role in transmission (28). The pooled estimated odds of being an infected contact among children compared to adults was 0.44 (95%CI 0.29–0.69). When restricted to household transmission studies alone, the pooled estimate was 0.19 (95% CI 0.10–0.36). While we see lower SARs among contacts who are children, this was only significant in the analysis that was restricted to confirmed secondary cases. When probable secondary cases were included the effect was no longer significant. This may reflect milder symptoms and a lower propensity for testing children, in which case previous estimates, with more stringent case definitions, in particular those relying on PCR confirmed cases alone would underestimate SARs in children. The review found no studies that reported SARs where children were the primary case. A review of household clusters by Zhu et al (2020) found that only 3 out of 31 household transmission clusters had a child as the index case, and suggested that children do not play a substantive role in transmission. Nevertheless, the low number of households with children as the index may be due to lower ascertainment in children if they are less likely to present with symptoms (29). However, recent evidence suggests that children carry higher levels of COVID-19 genetic material in their nose and throat than adults which would support our findings of a higher secondary attack rate among household contacts of children(30). Furthermore, a recent study from South Korea found that the highest proportion of positive household contacts by the age of the index case was among contacts of index cases aged 10–19 years of age (31). Nevertheless, the South Korean study did not identify whether index cases were the primary case therefore we do not know the direction of transmission.

Our study has a number of strengths: this is one of the largest COVID-19 household studies published to date and one of the only studies outside of Asia. Data was collected through direct patient interviews and high rates of follow-up were achieved with good data completeness, household contacts were actively followed up by local health protection teams on a daily basis to monitor symptoms. We identified point source case-secondary case pairs which allowed us to directly estimate the incubation period without having to model timing of infection. The study also has a number of limitations: test results were not available for some participants who developed symptoms, therefore we likely under-ascertained confirmed secondary cases. Furthermore, as with previous studies, testing was focussed on those who develop symptoms. Estimates of asymptomatic infection range from 4% to 41%, therefore we are likely to have missed asymptomatic cases (32). Furthermore, rates of asymptomatic infection appear to be highest in children, therefore we particularly underestimate secondary infection rates in children (29, 32). We are currently undertaking further analyses of household transmission incorporating swabbing of asymptomatics and serology which will provide a better understanding of true secondary infection rates and asymptomatic infection. We are also limited in our ability to draw any clear conclusions about transmission from children due to the small number of households with a child as a primary case.

Since the early stages of the pandemic, data from the FF100 study has been shared in real-time with independent modelling groups advising government and has informed policy making and public health management guidance. The high household SAR and the lack of transmission in a range of other settings highlight the importance of the household setting for onwards transmission. This emphasises the need for hygiene measures within the household and, where there are vulnerable members of the household, maintaining distancing within the household, in particular if a household member develops symptoms. While numbers are small, the high household SARs from paediatric primary cases suggest that reopening of schools needs careful monitoring for evidence of transmission from children in this setting.

## Data Availability

Applications for relevant anonymised data should be submitted to the Public Health England Office for Data Release: https://www.gov.uk/government/publications/accessing-public-health-england-data/about-the-phe-odr-and-accessing-data

